# The impact of hospital discharge on physical activity and sedentary behaviour following orthopaedic trauma: An interrupted time series analysis

**DOI:** 10.64898/2026.03.26.26349468

**Authors:** Asher G Kirk, Lara A Kimmel, Tyler Lane, Dot Dumuid, Christina L Ekegren

## Abstract

**Objectives:** To determine the impact of discharge home on physical activity and sedentary behaviour following orthopaedic trauma.

**Design:** Observational study.

**Setting:** Acute hospital.

**Participants:** Between October 2022 and January 2024, 31 adult orthopaedic trauma patients were recruited during hospital admission. Participants had either an isolated hip fracture or multi-trauma (i.e., a lower limb fracture, with an upper limb and/or spinal fracture).

**Interventions:** Participants wore two activity monitors (activPAL3 and ActiGraphGT3x) during the final days of an acute hospital admission and the first five days at home. An interrupted time series analysis evaluated changes physical activity variables during the hospital to home transition. Participants were analysed individually using mixed-effects linear regression allowing the intercept to vary by participant.

**Main outcome measures:** Primary outcome was daily steps; secondary outcomes included sedentary time and other activity measures.

**Results:** Daily steps (mean ± SD) were higher at home (4552.4 ±2639.5) compared to hospital (2597.8 ± 1450.8). Modelled results indicated a 27% increase in daily steps following hospital discharge (exp(β): 1.27, 95% CI: 1.01,1.59, p=0.039) and a sustained improvement at home. No significant differences were observed between hip fracture and multi-trauma participants.

**Conclusion:** Participants recovering from orthopaedic trauma showed a significant increase in daily step count upon discharge home from hospital, highlighting the positive impact of the home environment on activity levels. Further research is warranted to assess the effectiveness of interventions to improve activity levels in hospital (e.g., early intensive therapy) and at home (e.g., immediate home-based physiotherapy) in individuals following orthopaedic trauma.

## Introduction

The rising incidence of orthopaedic trauma presents a major public health and economic challenge ^[1]^. Orthopaedic trauma can consist of isolated injuries (e.g., hip fracture), or injury to multiple limbs (e.g., multi-trauma). Outcomes vary but often result in long term negative effects for the patient and community ^[2, 3]^. Hip fractures can lead to functional decline (i.e., loss of mobility and independence), higher long-term care needs ^[2, 4]^, and up to 30% mortality in the first 12-months post-injury ^[5]^. Compared to isolated injuries, multi-trauma is associated with high mortality rates, complications (e.g., infections), prolonged lengths of stay (LOS) and the need for inpatient rehabilitation ^[6–8]^.

Providing intensive allied health therapy in the acute hospital setting following trauma may improve long-term outcomes ^[9]^. For example, higher frequency and intensity of physiotherapy within the acute hospital has been shown to increase physical activity, improve physical function and reduce LOS in people recovering from orthopaedic surgery or trauma ^[10–13]^. Therefore, the hospital environment may facilitate engagement in physical activity, as long as adequate programs and sufficient staffing are in place. However, it is unclear whether physical activity is sustained post-discharge once intensive clinical interventions are withdrawn.

A recent systematic review found that, following hospitalisation, patients with diverse clinical presentations engage in more physical activity and less sedentary behaviour at home compared to both acute and subacute hospital settings ^[14]^. This increase in physical activity after the transition from hospital to home suggests that, despite the withdrawal of intensive therapy, the home environment itself may promote physical activity. For example, in patients recovering from a stroke, being discharged from hospital to home was associated with reduced sitting time and increased time spent walking ^[15]^. However, there is a lack of evidence investigating the objective change in activity levels following discharge home from hospital in people with orthopaedic trauma.

Understanding the change in activity levels from the hospital to home enables development of strategies in both environments to optimise recovery. Previous studies on changes in physical activity levels from hospital to home are limited by pre-post designs ^[15]^ which do not account for the natural course of recovery over time ^[16]^. In contrast, the interrupted time series (ITS) design evaluates time series data before and after an intervention is implemented (in this case, hospital discharge) to assess whether post-intervention trends significantly deviate from those predicted by pre-intervention trends ^[17, 18]^. As such, ITS offers a robust, quasi-experimental approach to derive causal inferences when randomisation is not possible ^[16]^. The objective of this study was to use an ITS study design to determine the impact of discharge home from hospital on physical activity and sedentary behaviour following orthopaedic trauma.

## Methods

### Participants

We recruited adult orthopaedic trauma patients during their acute hospital admission. The electronic medical record of patients admitted to orthopaedic or trauma units at [Blinded] was screened daily for eligibility. Inclusion criteria were: 1) age ≥18 years, 2) admitted for ≥24 hours, 3) predicted to be discharged home from hospital, and 4) experienced either an isolated hip fracture (managed operatively), or multi-trauma consisting of at least one lower limb fracture plus an upper limb or spinal fracture (confirmed by X-ray).

Exclusion criteria included: 1) pathological fracture, 2) severe traumatic brain injury, 3) spinal cord injury, 4) unable to walk owing to injury (e.g., bilaterally non-weight bearing lower limbs), 5) required physical assistance (other than a gait aid) to mobilise pre-morbidly, 6) resided in a residential aged care facility, 7) significant cognitive impairment. All participants provided written informed consent. [Blinded] Ethics Committee approved this study (Project number XXX/XX).

### Baseline measures

Participants’ age, sex, body mass index (BMI), admission date, injury details, orthopaedic/surgical management, weight-bearing orders, associated injuries and gait aid prescribed were collected from the Electronic Medical Record. The Clinical Frailty Scale (CFS), validated in people with hip fracture ^[19]^, was collected at baseline by an investigator [Blinded] during hospital admission.

### Outcomes

Physical activity and sedentary behaviour outcomes included steps per day (primary outcome), maximum steps completed in a single bout, sit to stand transitions, upright time, sedentary time, sedentary bouts of ≥ 30 minutes, light physical activity (LPA) and moderate-to-vigorous physical activity (MVPA).

Physical activity and sedentary behaviour data were continuously measured from the final days (up to seven days) of acute hospital admission to the first five days at home using the activPAL3™ triaxial accelerometer/inclinometer (Pal Technologies, Glasgow, Scotland) and the ActiGraph GT3x triaxial accelerometer (ActiGraph Corp, Pensacola, Florida) [21]. The placement and use of each activity monitor were guided by a recent study investigating their criterion validity in individuals hospitalised with lower limb fractures, including those who were managed partial- and non-weight bearing ^[20]^. The activPAL was worn on the anterior middle third of the thigh on the non-injured leg, secured with Tegaderm™ (3M Healthcare, St. Paul, MN), and participants were instructed to wear it for 24 hours/day throughout the monitoring period. The ActiGraph, secured to the unaffected ankle with an elastic band, was worn for 24 hours/day, except for showering. The activPAL was used to assess sedentary time (sitting/lying), sit-to-stand transitions, and upright time (standing and walking). The ActiGraph was used to measures steps, LPA and MVPA.

### Data processing

Manufacturer software was used to download activPAL (activPAL3^TM^ v7.2.32) data in 1-second epochs and ActiGraph (ActiLife 6 v6.13.4) data in 10-second epochs. The activPAL and ActiGraph both had a sampling frequency of 30Hz, and the ActiGraph data were downloaded using the low frequency extension filter (LFEF), as recommended for slow gait speeds ^[21]^. Data were included only for complete days of monitoring (i.e., 24 hours), excluding the days of recruitment and hospital discharge. We excluded the day of hospital discharge as it was difficult to account for time spent in either hospital or home settings (e.g., transport time returning home). Therefore, the following day was considered day 1 at home.

### Data analysis

All participants provided physical activity data in both the hospital and home settings. In hospital, 76% of participants had between two and four valid full wear days, whereas at home the majority of participants (74%) had five or six valid full wear days (Appendix A).

Therefore, we included physical activity data from three days pre- and five days post-discharge home. To account for any missing physical activity data on these days, we conducted random forest multiple imputation using the missRanger R package ^[22]^, which fills in missing values by generating plausible values derived from distributions and relationships among observed variables in the data set. Variables included in the prediction model included each physical activity and sedentary behaviour outcome, participant identifier, sex, age, injury type (hip or multi-fracture), whether the participant had surgery, time since surgery, holiday/weekend day, and analytical variables (time, trend, intervention [0/1]; described below). Twenty imputations were generated; each was analysed separately and results pooled according to Rubin’s rules ^[23]^.

Descriptive statistics were used to describe participant characteristics, baseline measures and imputed physical activity/sedentary behaviour outcomes (Stata version 17, StataCorp). The ITS analysis was completed in R using the imputed datasets ^[24]^. The intervention was defined as a binary variable distinguishing between the days pre- and post-transition home to model the change to the intercept. Trend was defined as a continuous variable, set at zero pre-intervention and counting up from one each day sequentially post-intervention to model the change to the slope. Time was a continuous variable, counting each day sequentially from the start of the time series, including three days pre- and five days post-transition home (the fourth day on which the transition occurred was omitted), to model the secular (or pre-intervention) trend.

The ITS modelling was carried out using mixed-effects linear regression, allowing intercepts to vary by participant. Additional analyses included an interaction term between the intervention (discharge home) and injury type to determine whether effects varied between hip fracture and multi-trauma patients. Interaction model results are reported as the percentage change to the slope and intercept in hip fracture patients, and percentage point differences in changes in multi-trauma patients. For instance, if the intercept change for steps in hip fracture patients was 10 and the change in multi-trauma patients was -3, these would be interpreted as a 10% increase in steps among hip fracture patients and a 7% increase in steps among multi-trauma patients (3 percentage points lower than the change in hip fracture patients). Analyses adjusted for confounders shown to be associated with physical activity outcomes in trauma populations, including age, sex, injury type (hip fracture vs multi-trauma), operative vs non-operative management, time since surgery, and weekend or public holiday ^[25–28]^.

## Results

### Participant characteristics

Thirty-one participants were recruited between October 2022 and January 2024 (Figure 1). The median (Interquartile range [IQR]) age was 67.0 (57.0-79.5) years, and 58% of participants were female, with a similar number of hip fracture (49%) and multi-trauma (51%) injury types (Table 1). Most participants had non-transport related injuries (68%), with a median (IQR) acute hospital LOS of 6.1 (5.7-7.8) days. Few participants were under non-weight bearing orders (i.e. hopping) (19%). All participants were referred for ongoing physiotherapy following hospital discharge. However, only one participant commenced physiotherapy within the home monitoring period.

**Figure 1.**
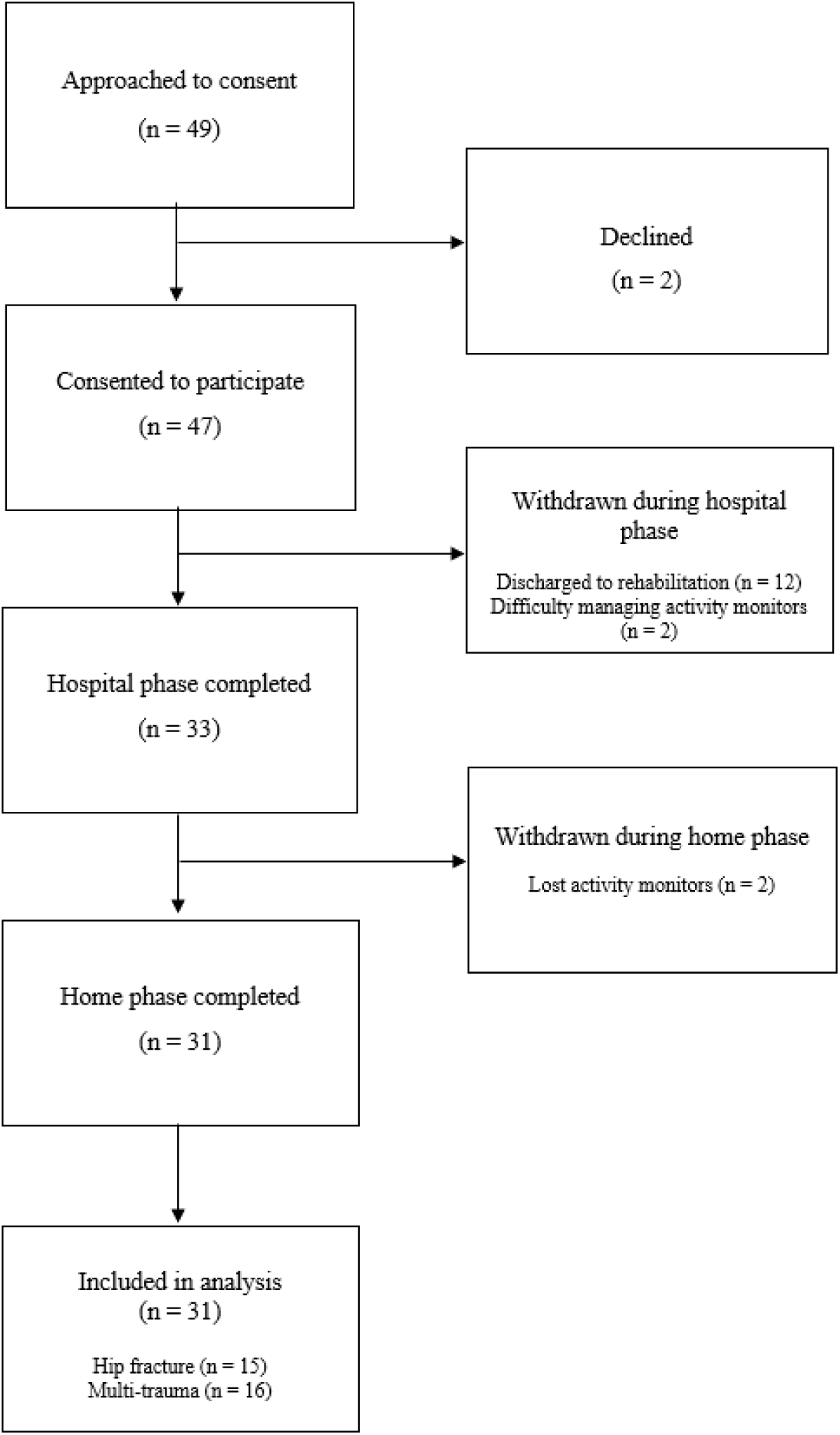
Flow of participants through the study.

**Table 1.** Participant characteristics (n=31)

| **Characteristic** | **n (%) or median (IQR)** |
| --- | --- |
| Age (years) | 67.0 (57.0-79.5) |
| Age group |  |
| <65 years | 12 (39%) |
| ≥65 years | 19 (61%) |
| Female sex | 18 (58%) |
| Type of injury |  |
| Hip fracture | 15 (48%) |
| Multi-trauma | 16 (51%) |
| Mechanism of injury |  |
| Low fall | 12 (39%) |
| Transport | 10 (32%) |
| Other | 9 (29%) |
| LOS in hospital (days) | 6.1 (5.7-7.8) |
| Underwent surgery | 25 (81%) |
| Duration from surgically complete to hospital discharge (days)* | 5.2 (4.3-7.1) |
| Clinical Frailty Scale |  |
| 1 □ | 13 (42%) |
| □ 2-4 □ | 18 (58%) |
| Lower limb weight bearing status □ on discharge □ |  |
| Weight bearing (WBAT, PrWB, PWB) □ □ | 25 (81%) |
| Non or touch weight bearing □ □ | 6 (19%) |
| Gait aid on discharge |  |
| Crutches (forearm or axillary) | 14 (45%) |
| Frame (GF/PUF/2WF/4WF) | 17 (55%) |
| Follow-up physiotherapy (post hospital discharge) |  |
| Home-based rehabilitation | 17 (55%) |
| Community-based rehabilitation | 14 (45%) |

IQR – interquartile range; LOS – length of stay; WBAT – weight bearing as tolerated; PrWB – protected weight bearing; PWB – partial weight bearing; GF – gutter frame; PUF – pick up frame; 2WF – two wheeled frame; 4WF – four wheeled frame. \*for participants who underwent surgery (n=25).

### Physical activity and sedentary behaviour levels in hospital and home settings

The mean (SD) activity levels for each outcome within the hospital and home setting for all participants are detailed in Table 2. Overall, participants engaged in more physical activity and less sedentary behaviour at home compared to hospital.

**Table 2:** Mean (SD) physical activity and sedentary behaviour levels in hospital and at home (using first imputed dataset to account for missing data)

| **Activity type** | **Hospital**<br>**Mean (SD)** | **Home**<br>**Mean (SD)** | **Overall**<br>**Mean (SD)** |
| --- | --- | --- | --- |
| Steps per day (n) | 2597 (1450) | 4552 (2639) | 3907 (2488) |
| Maximum steps completed in a single bout (n) | 14 (8) | 17 (8) | 16 (8) |
| Sit to stand transitions (n) | 18 (11) | 30 (14) | 26 (14) |
| Sedentary time (mins) | 860.7<br>(266.2) | 777.6<br>(232.5) | 805.0<br>(246.6) |
| Upright time (mins) | 65.6 (63.0) | 131.6 (78.1) | 109.8 (79.6) |
| Sedentary time spent in bouts >30 mins (mins) | 794.0<br>(279.0) | 637.7<br>(277.3) | 689.3<br>(286.8) |
| Light physical activity (mins) | 85.8 (48.4) | 112.9 (51.6) | 103.9 (52.0) |
| Moderate-to-vigorous physical activity (mins) | 3.0 (3.8) | 12.4 (17.1) | 9.3 (14.8) |

### Interrupted time series analysis

The original intention was to report effects of both the change in intercept and change in trend to reflect the intervention – hospital discharge – on outcomes. However, in many cases the pre-discharge secular trend increase was steep, indicating unsustainable progress beyond a few days, so that in many cases the post-discharge trend coefficient was negative, which was at odds with what the plotted time series indicated. Therefore, for each outcome, we have reported the coefficients for the change in intercept only and have interpreted trend effects qualitatively based on patterns visible in time series plots. However, the coefficients for both the change in intercept and trend are provided in Appendix B.

For the primary outcome (steps per day), there was a 27% increase in the change in the intercept following hospital discharge, based on modelled and plotted results (exp(β): 1.27, 95% CI: 1.01, 1.59, p=0.039) (Appendix B). Notably, the change in the trend based on plotted results showed that step count remained relatively stable within the home environment post-discharge (Figure 2).

**Figure 2.**
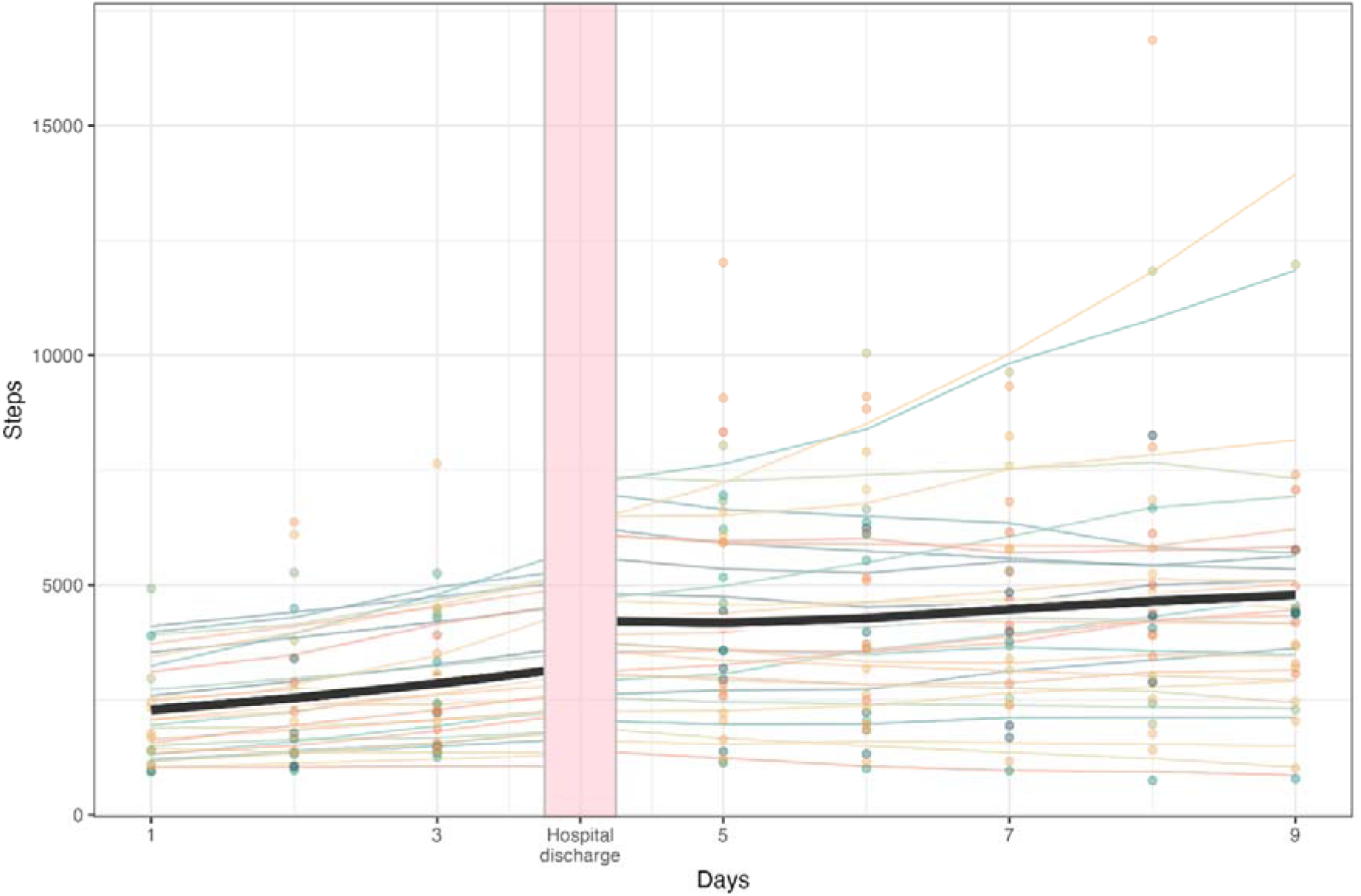
Interrupted time series plots for steps per day (n=31) Thick black line corresponds to the outcome average across participants from the pooled regression model; faded thin lines correspond to individually-fit data from the pooled regression model, colour-coded by participant; faded points correspond to actual outcome data (with missing points supplemented by data from imputation #1)

For secondary outcomes, there were varying effects of hospital discharge. A 20% increase in maximum steps completed in a single bout (exp(β): 1.20, 95% CI: 1.01, 1.44, p=0.048) was seen (Appendix B). These effects were sustained within the home environment (Figures 3A and 4D). There was a non-significant increase in sit to stand transitions (exp(β): 1.23, 95% CI: 0.97, 1.57, p=0.090) and upright time (exp(β): 1.41, 95% CI: 0.98, 2.03, p=0.066) (Appendix B). There were no meaningful changes post-discharge for sedentary time, sedentary time spent in bouts of >30 minutes, and LPA. Plotted results indicated that secondary outcomes remained relatively stable in the home environment, except sit to stand transitions, for which the trend could be interpreted as a gradual improvement post-discharge (Figure 3B). Very few participants engaged in any MVPA, meaning this outcome was excluded from analysis.

**Figure 3.**
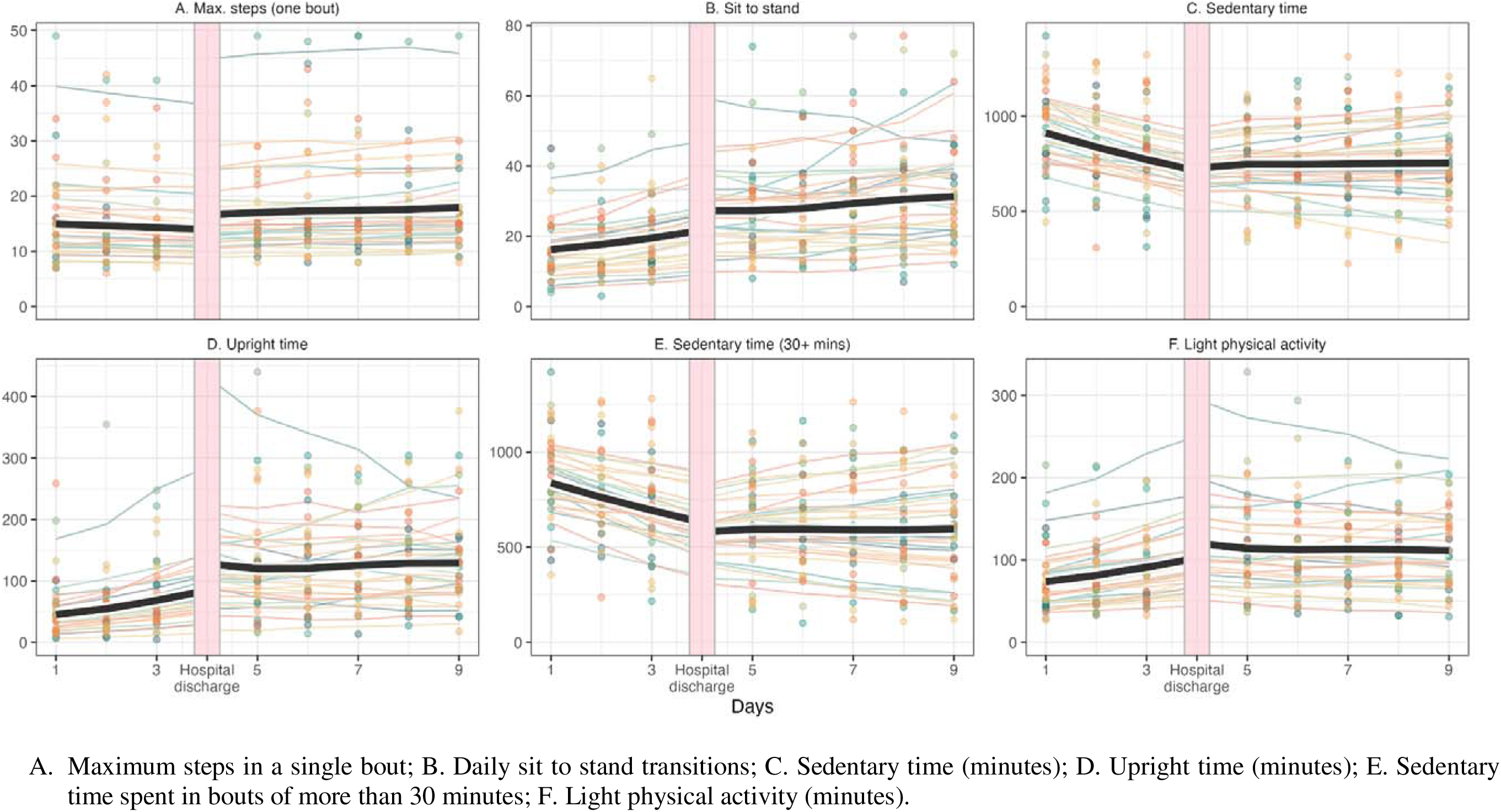
Interrupted time series plots for secondary outcomes (n=31) Thick black line corresponds to the outcome average across participants from the pooled regression model; faded thin lines correspond to individually-fit data from the pooled regression model, colour-coded by participant; faded points correspond to actual outcome data (with missing points supplemented by data from imputation #1)

**Figure 4.**
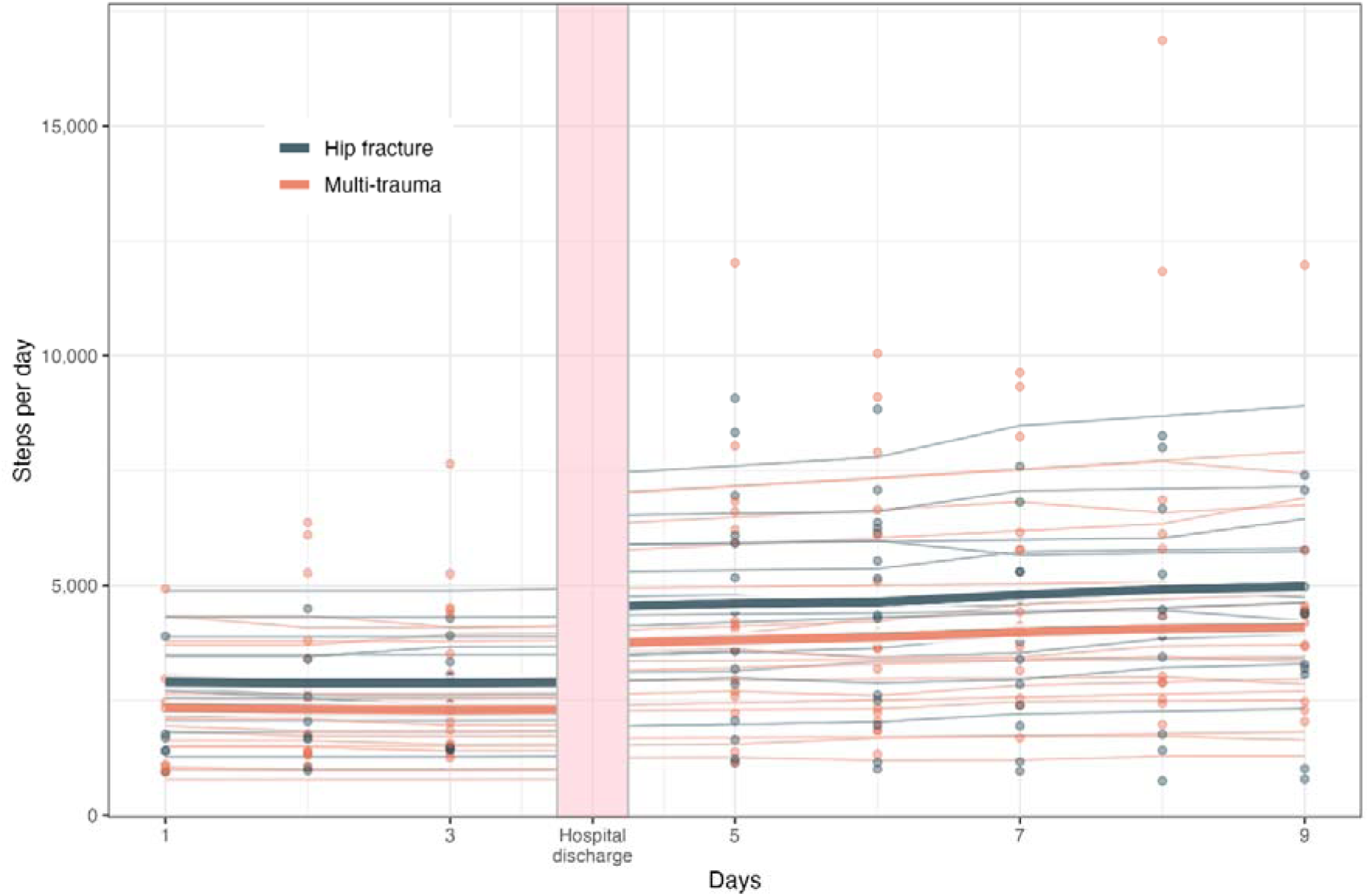
Interrupted time series plots for steps per day comparing participants with hip fracture (n=15) and multi-trauma (n=16) Thick lines corresponds to the outcome average across participants from the pooled regression model; faded thin lines correspond to individually-fit data from the pooled regression model, colour-coded by participant; faded points correspond to actual outcome data (with missing points supplemented by data from imputation #1)

### Subgroup analyses (hip fracture vs. multi-trauma)

No detectable difference was seen in steps per day between participants with hip fracture and multi-trauma for either the post-discharge change in intercept (p=0.989) or trend (p=0.654). The model suggested steps per day increased by 27% (exp(β): 1.27, 95% CI: 0.96, 1.68, p=0.099) among participants with hip fractures; the effect in participants with multi-trauma was marginal and not significantly different (exp(β): 1.01, 95%CI: 0.75, 1.35, p=0.977) or no change compared to participants with hip fracture) (Appendix C). The change in the trend based on plotted results demonstrated that steps per day remained relatively stable in both groups within the home environment (Figure 4).

There were no detectable differences in the effect of transition to home on any of the secondary outcomes between patients with hip fractures and multi-trauma (Appendix C). The change in the trend based on plotted results demonstrated that all secondary outcomes remained relatively stable in the home environment (Figure 5A to 5F).

**Figure 5.**
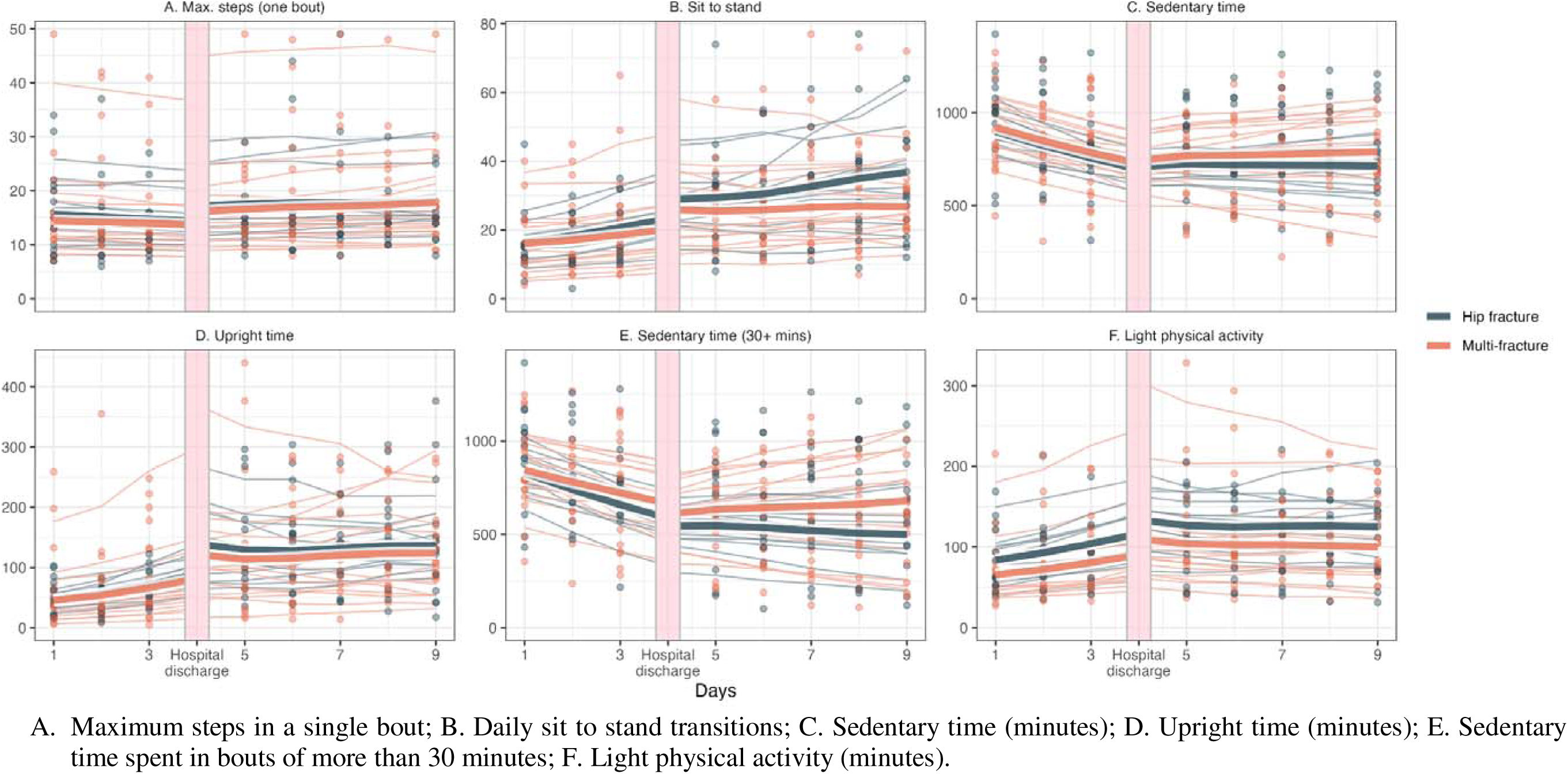
Interrupted time series plots for secondary outcomes comparing participants with hip fracture (n=15) and multi-trauma (n=16) Thick lines corresponds to the outcome average across participants from the pooled regression model; faded thin lines correspond to individually-fit data from the pooled regression model, colour-coded by participant; faded points correspond to actual outcome data (with missing points supplemented by data from imputation #1)

## Discussion

To our knowledge, this study is the first to examine the impact of being discharged home from hospital on physical activity and sedentary behaviour in people recovering from orthopaedic trauma. We found that participants engaged in more physical activity and less sedentary behaviour at home compared to the acute hospital setting, suggesting that being discharged home can immediately improve patients’ activity levels following orthopaedic trauma.

Participants consistently engaged in a higher daily step count at home compared to hospital, aligning with previous findings ^[29]^. Higher physical activity post-hospital discharge is expected, given the natural course of recovery. However, by accounting for the secular trend in our analysis, the significant increase in daily steps upon returning home suggests additional factors influencing physical activity beyond natural recovery. Patients recovering from fractures have reported there is often no reason to be physically active or engage in activities of daily living (ADLs) during hospitalisation, aside from toileting ^[30]^. In contrast, upon discharge home, participants are required to engage in ADLs and subsequent physical activity. Furthermore, policies and clinical oversight in hospital often reinforce a risk-reductive approach, which can inadvertently reduce opportunities for patients to safely engage in activity, therefore potentially limiting independence ^[31]^. Patients with orthopaedic trauma sometimes perceive a lack of permission from hospital staff to remain active ^[30]^, perhaps contributing to the observed increase in physical activity upon returning home, where hospital constraints no longer apply. To support more activity within hospitals, policy changes may be required to harness safe physical activity behaviours and promote independent functional activities.

While overall physical activity levels increased immediately upon discharge home, participants experienced a less rapid rate of improvement – or sustained improvement – at home, compared to in hospital. Potentially, participants just improved more quickly in the earlier days following injury/surgery. However, this more rapid rate of improvement in hospital could also be attributed to the comprehensive care provided during hospitalisation (including at least daily physiotherapy), whereas at home, support was limited, with only one participant engaging in physiotherapy during the monitoring period. Implementing a multi-disciplinary, individualised, home-based rehabilitation program after hospital discharge has been shown to enhance patient independence and physical activity ^[32, 33]^. Furthermore, given patients have reported uncertainty about the importance of physical activity for recovery post-trauma ^[30]^, education, advice and the use of technology (i.e., wearable devices/health apps) for goal setting and progress monitoring are important strategies for improving the rate of improvement in activity levels post-discharge ^[34, 35]^.

Activity patterns observed in participants with hip fracture and those with multi-trauma were comparable during and after hospitalisation. Several factors may contribute to the absence of a significant difference between the two injury groups. Given that participants were monitored during the final days of their acute hospital admission, it is likely that both groups reached a similar functional level in preparation for discharge home, potentially explaining the similar rate of improvement at home. However, the relatively small sample size and the brief monitoring period limited the statistical power to detect differences between participants within the two groups. Additionally, the heterogeneity among the multi-trauma participants likely introduced greater outcome variability, complicating direct comparison with the more homogeneous group of individuals with hip fractures.

There are some limitations to consider. First, the sample size was relatively small. However, this was countered by the number of observations for each participant; when maximised with multiple imputation to account for missing observations, this resulted in 31 individual time series with 7 observations each, or 217 data points. Second, the monitoring periods in the hospital and home settings were limited to three and five days, respectively. While this duration may appear brief, a minimum of four valid wear days has been recommended for reliable estimation of physical activity in free-living conditions ^[36]^. Evidence establishing the minimum wear time required for accurate quantification of physical activity in hospital settings remains limited. However, the relatively stable nature of the hospital environment suggests a three-day monitoring period may be sufficient. Third, although the median age of participants was 67 years, the study did not include any frail individuals. Frail trauma patients have been shown to have a low level of physical activity during hospitalisation and often experience adverse outcomes post discharge ^[37]^. Therefore, future research is needed to investigate the influence of frailty on physical activity during the transition from hospital to home in people with orthopaedic trauma. Finally, as this was a single-site study, the findings may not be generalisable to other institutions with differing allied health support and home-based rehabilitation services.

A strength of this study is that, to our knowledge, it is the first to continuously monitor activity levels during the transition from hospital to home in individuals recovering from orthopaedic trauma. The use of random forest multiple imputation and pooling of results according to Rubin’s rules applied a sophisticated approach towards managing missing data and mitigating any potential bias, while also maximising statistical power. Additionally, our rigorous method for evaluating longitudinal data, the ITS, enabled assessment of whether discharge from hospital causally affected physical activity and sedentary behaviour outcomes.

## Conclusions and implications

Following orthopaedic trauma, participants demonstrated a significant increase in daily step count at home following hospital discharge, highlighting the immediate benefit of home discharge on activity levels. Overall recovery of activity levels remained relatively stable in the home setting suggesting ongoing support is necessary to continually improve activity levels post discharge. Further research is warranted to explore the impact of interventions (e.g., early and intensive acute hospital therapy, immediate home-based physiotherapy, goal setting with wearable devices) on enhancing activity levels both during hospitalisation and post-discharge in individuals recovering from orthopaedic trauma. Additionally, studies assessing the impact of physical activity recovery on other key patient outcomes (e.g., quality of life) are needed.

## Data Availability

All data produced in the present study are available upon reasonable request to the authors

---

**Appendix A.** Number and percentage of participants according to the number of valid days (i.e., 24-hour wear) in each setting.

| **Number of valid days** | **Hospital, n (%)** | **Home, n (%)** |
| --- | --- | --- |
| 2 | 7 (22%) | 3 (10%) |
| 3 | 10 (32%) | 1 (3%) |
| 4 | 7 (22%) | 4 (13%) |
| 5 | 2 (6.5%) | 19 (61%) |
| 6 | 3 (10%) | 4 (13%) |
| 7 | 2 (6.5%) | 0 (0%) |

**Appendix B.** Coefficients for the change in intercept and change in trend for all outcomes (primary effects)

| Outcome | Term | exp($\beta$) | 95% CI | p |
| --- | --- | --- | --- | --- |
| Steps per day | Change in intercept | 1.27 | 1.01 to 1.59 | 0.039 |
|  | Change in trend | 0.9 | 0.81 to 1.00 | 0.054 |
| Max steps completed in a single bout | Change in intercept | 1.2 | 1.00 to 1.44 | 0.048 |
|  | Change in trend | 1.04 | 0.95 to 1.12 | 0.394 |
| Sit to stand transitions | Change in intercept | 1.23 | 0.97 to 1.57 | 0.090 |
|  | Change in trend | 0.92 | 0.82 to 1.03 | 0.166 |
| Sedentary time | Change in intercept | 1.05 | 0.87 to 1.27 | 0.631 |
|  | Change in trend | 1.08 | 0.99 to 1.18 | 0.073 |
| Upright time | Change in intercept | 1.41 | 0.98 to 2.03 | 0.066 |
|  | Change in trend | 0.8 | 0.68 to 0.94 | 0.009 |
| Sedentary time spent in bouts >30 mins □ | Change in intercept | 0.95 | 0.72 to 1.24 | 0.691 |
|  | Change in trend | 1.09 | 0.97 to 1.23 | 0.150 |
| Light physical activity | Change in intercept | 1.13 | 0.93 to 1.38 | 0.229 |
|  | Change in trend | 0.88 | 0.80 to 0.96 | 0.007 |

\*moderate-to-vigorous physical activity was not included due to very limited data for this outcome
exp( $\beta$ ) - where “ $\beta$ ” refers to the beta coefficient and “exp()” indicates exponentiation to give the multiplicative change in the outcome; when subtracted by 1 and multiplied by 100, this corresponds to percent change in the outcome

**Appendix C.**
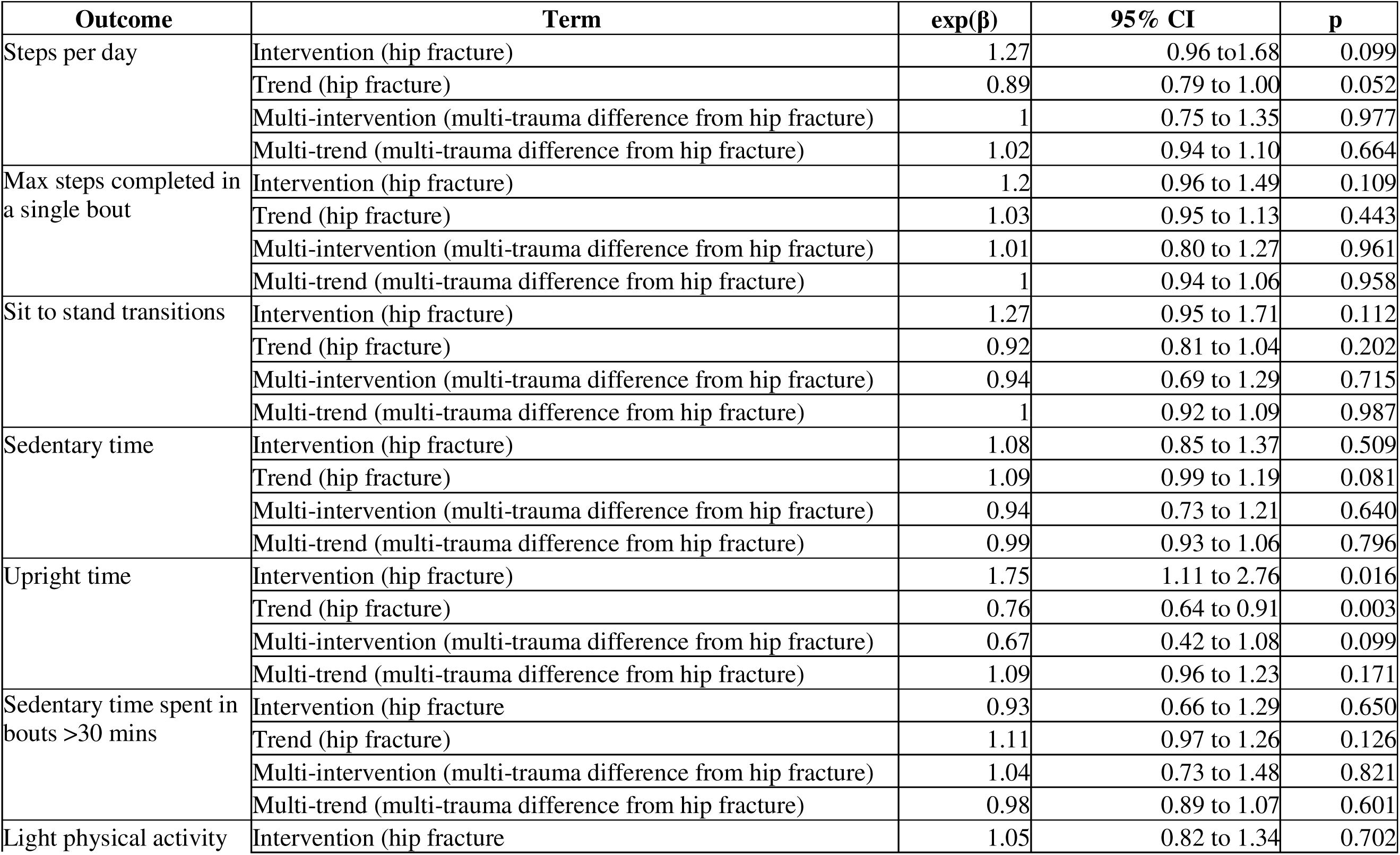
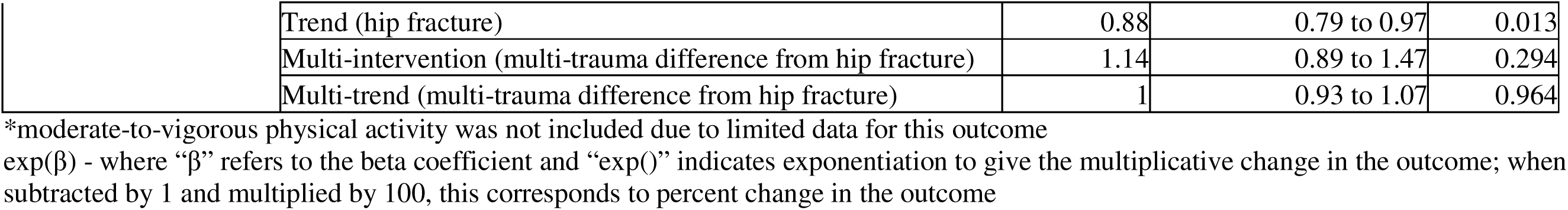
Coefficients for the change in intercept and change in trend for all outcomes (injury interaction effects -hip fracture vs. multi-trauma)

## References

1. Wu, A.-M., et al., Global, regional, and national burden of bone fractures in 204 countries and territories, 1990&#x2013;2019: a systematic analysis from the Global Burden of Disease Study 2019. The Lancet Healthy Longevity, 2021. 2(9): p. e580–e592.

2. Dyer, S.M., et al., A critical review of the long-term disability outcomes following hip fracture. BMC Geriatr, 2016. 16(1): p. 158.

3. O’Hara, N.N., et al., The socioeconomic impact of orthopaedic trauma: A systematic review and meta-analysis. PLoS One, 2020. 15(1): p. e0227907.

4. Leal, J., et al., Impact of hip fracture on hospital care costs: a population-based study. Osteoporos Int, 2016. 27(2): p. 549–58.

5. Downey, C., M. Kelly, and J.F. Quinlan, Changing trends in the mortality rate at 1-year post hip fracture - a systematic review. World J Orthop, 2019. 10(3): p. 166–175.

6. Anghele, M., et al., Negative Factors Influencing Multiple-Trauma Patients. Clinics and Practice, 2024. 14(4): p. 1562–1570.

7. Dlj, M., et al., Concurrent upper limb and hip fracture in the elderly. Injury, 2020. 51(4): p. 1025–1030.

8. Pape, H.C., et al., Timing of major fracture care in polytrauma patients - An update on principles, parameters and strategies for 2020. Injury, 2019. 50(10): p. 1656–1670.

9. Kimmel, L., et al., Outcomes following intensive allied health therapy in the acute hospital for trauma patients. Injury, 2024: p. 111942.

10. Calthorpe, S., et al., An intensive physiotherapy program improves mobility for trauma patients. J Trauma Acute Care Surg, 2014. 76(1): p. 101–6.

11. Cogan, A.M., et al., Association of Length of Stay, Recovery Rate, and Therapy Time per Day With Functional Outcomes After Hip Fracture Surgery. JAMA Netw Open, 2020. 3(1): p. e1919672.

12. Kimmel, L.A., et al., HIP4Hips (High Intensity Physiotherapy for Hip fractures in the acute hospital setting): a randomised controlled trial. Med J Aust, 2016. 205(2): p. 73–8.

13. Siminiuc, D., et al., Rehabilitation after surgery for hip fracture – the impact of prompt, frequent and mobilisation-focused physiotherapy on discharge outcomes: an observational cohort study. BMC Geriatrics, 2024. 24(1): p. 629.

14. Kirk, A.G., et al., Levels of Physical Activity and Sedentary Behavior During and After Hospitalization: A Systematic Review. Arch Phys Med Rehabil, 2021. 102(7): p. 1368–1378.

15. Simpson, D.B., et al., Go Home, Sit Less: The Impact of Home Versus Hospital Rehabilitation Environment on Activity Levels of Stroke Survivors. Arch Phys Med Rehabil, 2018. 99(11): p. 2216–2221 e1.

16. Shadish, W.R., T.D. Cook, and D.T. Campbell, Experimental and quasi-experimental designs for generalized causal inference. Experimental and quasi-experimental designs for generalized causal inference. 2002, Boston, MA, US: Houghton, Mifflin and Company. xxi, 623–xxi, 623.

17. Soumerai, S.B., D. Starr, and S.R. Majumdar, How Do You Know Which Health Care Effectiveness Research You Can Trust? A Guide to Study Design for the Perplexed. Prev Chronic Dis, 2015. 12: p. E101.

18. Wagner, A.K., et al., Segmented regression analysis of interrupted time series studies in medication use research. J Clin Pharm Ther, 2002. 27(4): p. 299–309.

19. Kay, R.S., et al., The Clinical Frailty Scale can be used retrospectively to assess the frailty of patients with hip fracture: a validation study. European Geriatric Medicine, 2022. 13(5): p. 1101–1107.

20. Kirk, A.G., et al., Validity of the activPAL and ActiGraph for measuring sitting time and steps in hospitalised orthopaedic patients with altered weight bearing. Disabil Rehabil, 2022: p. 1–9.

21. Feito, Y., H.R. Garner, and D.R. Bassett, Evaluation of ActiGraph’s low-frequency filter in laboratory and free-living environments. Med Sci Sports Exerc, 2015. 47(1): p. 211–7.

22. Mayer, M., missRanger: Fast imputation of missing values. R package version, 2019. 2(0).

23. Rubin, D.B., Multiple imputation for nonresponse in surveys. Vol. 81. 2004: John Wiley & Sons.

24. Team, R.C., R: A Language and Environment for Statistical Computing. 2024, R Foundation for Statistical Computing: Vienna, Austria.

25. Liao, Y.H., et al., Gender differences in the association between physical activity and health-related quality of life among community-dwelling elders. Aging Clin Exp Res, 2021. 33(4): p. 901–908.

26. Ekegren, C.L., et al., Physical Activity and Sedentary Behavior 6 Months After Musculoskeletal Trauma: What Factors Predict Recovery? Phys Ther, 2020. 100(2): p. 332–345.

27. Ekegren, C.L., et al., Physical Activity and Sedentary Behavior Subsequent to Serious Orthopedic Injury: A Systematic Review. Arch Phys Med Rehabil, 2018. 99(1): p. 164–177 e6.

28. Kirk, A.G., et al., Physical Activity in the Acute Hospital Following Elective Lower Limb Joint Arthroplasty. New Zealand Journal of Physiotherapy, 2022. 50(1).

29. Baldwin, C., et al., Accelerometry Shows Inpatients With Acute Medical or Surgical Conditions Spend Little Time Upright and Are Highly Sedentary: Systematic Review. Phys Ther, 2017. 97(11): p. 1044–1065.

30. Kirk, A.G., et al., The influence of hospital and home environments on physical activity and sedentary behaviour: Perceptions of people recovering from fractures. Injury, 2024. 55(4).

31. Growdon, M.E., R.I. Shorr, and S.K. Inouye, The Tension Between Promoting Mobility and Preventing Falls in the Hospital. JAMA Intern Med, 2017. 177(6): p. 759–760.

32. Fairhall, N.J., et al., Interventions for improving mobility after hip fracture surgery in adults. Cochrane Database Syst Rev, 2022. 9(9): p. Cd001704.

33. Zidén, L., K. Frändin, and M. Kreuter, Home rehabilitation after hip fracture. A randomized controlled study on balance confidence, physical function and everyday activities. Clin Rehabil, 2008. 22(12): p. 1019–33.

34. Aarden, J.J., et al., Recommendations for an exercise intervention and core outcome set for older patients after hospital discharge: Results of an international Delphi study. PLoS One, 2023. 18(3): p. e0283545.

35. Taylor, N.F., et al., Behaviour change interventions to increase physical activity in hospitalised patients: a systematic review, meta-analysis and meta-regression. Age Ageing, 2022. 51(1).

36. Trost, S.G., K.L. McIver, and R. Pate, Conducting Accelerometer-Based Activity Assessments in Field-Based Research. Medicine & Science in Sports & Exercise, 2005. 37(11): p. S531–S543.

37. Zhao, F., et al., The impact of frailty on posttraumatic outcomes in older trauma patients: A systematic review and meta-analysis. J Trauma Acute Care Surg, 2020. 88(4): p. 546–554.

